# Forecasting the Spreading Trajectory of the COVID-19 Pandemic

**DOI:** 10.1101/2021.03.26.21254429

**Authors:** Baolian Cheng, Yi-Ming Wang

## Abstract

Predictively forecasting future developments for the spread of the COVID-19 pandemic is extremely challenging. A recently published logistic mathematic model has achieved good predictions for infections weeks ahead. In this short communication, we summarize the Logistic spread model, which describes the dynamics of the pandemic evolution and the impacts of people social behavior in fighting against the pandemic. The new pandemic model has two parameters (i.e., transmission rate γ and social distancing d) to be calibrated to the data from the pandemic regions in the early stage of the outbreak while the social distancing is put in place. The model is capable to make early predictions about the spreading trajectory in the communities of any size (countries, states, counties and cities) including the total infections, the date of peak daily infections and the date of the infections reaching a plateau if the testing is sufficient. The results are in good agreement with data and have important applications for ongoing outbreaks and similar infectious disease pandemics in the future.

## Introduction

The highly contagious and asymptomatic transmission nature of the novel coronavirus has led to the explosive spreading of the COVID-19 pandemic [1–3] and has drastically affected the world economy. Unlike past flu seasons in the past, without any intervention, the infection of COVID-19 will follow a natural exponential growth path in time until most of the population is infected [4]. Social distancing plays a critical role in reducing the spread of the pandemic and flattens the infection curve. Many flu-based pandemic models [5–9] have been used to model the COVID-19 pandemic and severely missed predicting the rate of spread and peak time of new infections during the first wave in 2020 due to the lack of a social distancing requirement. The recently published logistic spread model [10, 11] addresses this issue and has been successfully applied to various of pandemic regions from cities to counties to states or countries of any size. The model predicts the number of total infections, daily infection rate, time of peak new daily infections, and time of infection to reach the 96% of the total infections of the pandemic weeks ahead, and the predictions are in good agreement with the observed data. The results provide possible guidance for policy makers on when and how to reopen the economy for pandemic regions.

### Model

A new logistic model [10, 11] is recently published to describe the spread of the COVID-19 pandemic under the regulation of social distancing. The model provides analytic solutions for the trajectory of the pandemic spread (daily and cumulative infections and the time of peak daily infections) weeks in advance. The total infected population *P(t)* at a given time in the model is described by function,

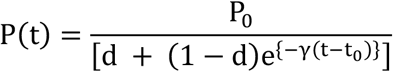

where P_0_ is the number of infected people at time t_0_, γ is the community transmission rate, and d is the social distancing level. Parameter d has values between d_min_ (no isolation or social distancing) and 1 (complete isolation or infinite social distancing) depending on people’s social behavior. The minimum value of d is determined by d_min_ = P_0_/[(1-η)P_max_], here η represents the fraction of the population who are naturally immune to the virus, and P_max_ is the total population of the community. The time derivative of P(t), dP(t)/dt, describes the daily new infections. In the model, increased social distancing with time would reduce the daily infection rate. On the other hand, relaxing social distancing over time will increase the daily infections. The parameters g and d are calibrated to data from a given community at the time when social distancing was put into effect.

The meaning of social distancing in the model is extended to include physical isolation, sheltering in place, staying at home, wearing face masks in public, washing hands, and restricting group-gathering size and frequency. If no social distancing is implemented, d = d_min_ ≈ 0, then, 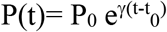 and 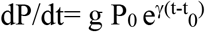, both the total and daily infections grow exponentially with time until the number of susceptible individuals is depleted, or nearly all people, (1-η) P_max_, have been infected. But if all the infected people are clearly traced, identified, and completely isolated, d=1, then, P(t)=P_0_ and dP/dt =0, there would be no spread at all and hence no pandemic.

## Results

The new Logistic model has been applied to numbers of countries and states as well as counties in the United States [10, 11, 13, 14] during the first wave in 2020. The model successfully predicts the peak time of daily infections, the infection doubling time, daily infection rate and the cumulative infections in time. As examples, Fig. 1 and Fig. 2, respectively, display the model predictions for the daily and total infections in time in the South Korea and the Germany compared with the real observed infection data [16, 17] during the first wave.

**Fig. 1.**
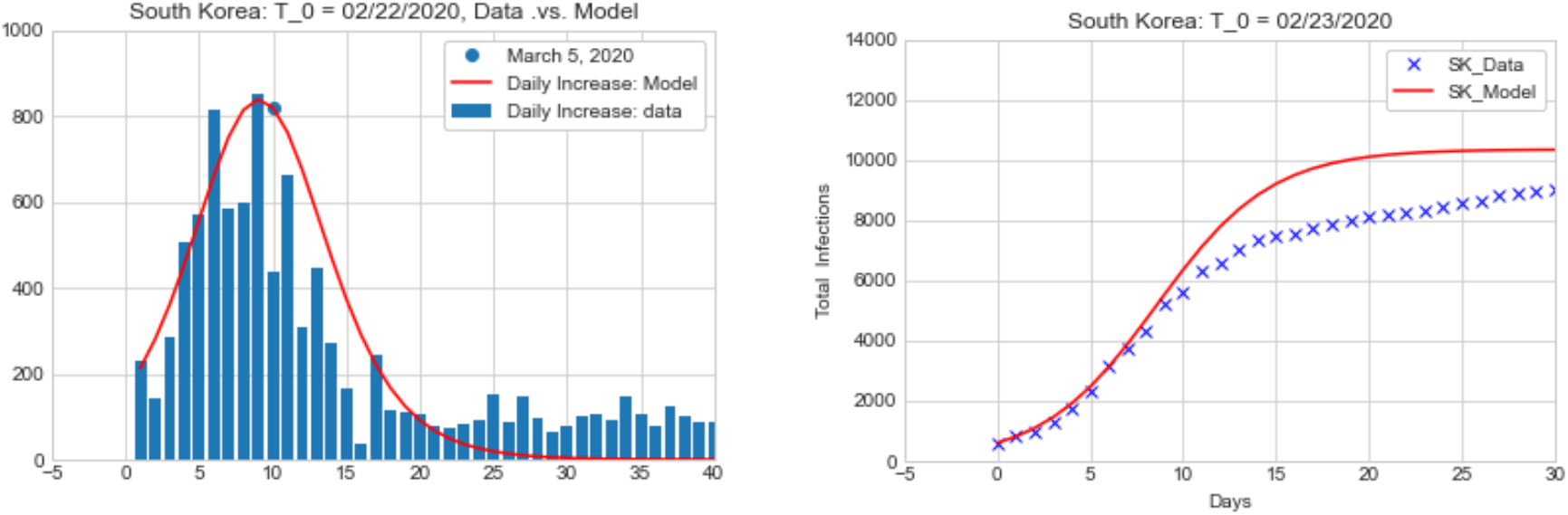
Model predictions for the daily infections (left) and total infections (right) in time vs. real infection data in the South Korea. The initial time t_0_=02/23/2020, P_0_=602, γ=0.325 and d=0.0778.

**Fig. 2.**
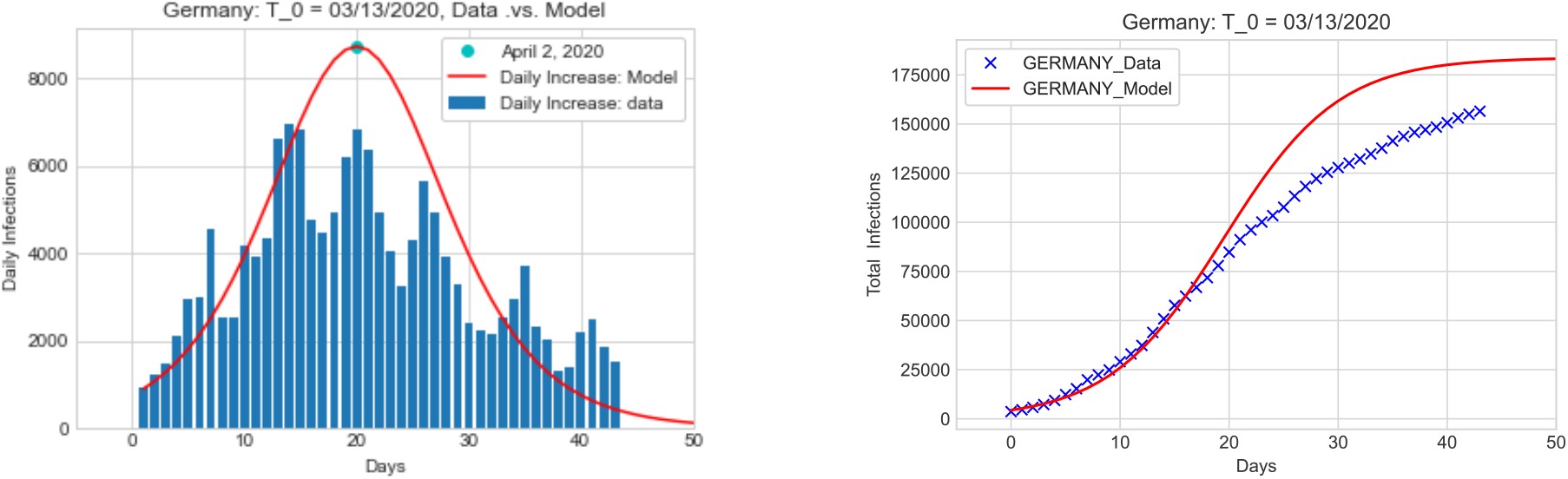
Model predictions for the daily infections (left) and total infections (right) in time vs. real infection data in the Germany. The initial time t_0_=03/13/2020, P_0_=3675, γ=0.2 and d=0.075.

The results show that the model predictions are in good agreement with data, particularly for countries with excellent testing capability. The accuracy of the model predictions strongly depends on the testing capacity or the positivity rate (number of positive cases divided by the number of people tested) in the pandemic regions [10]. The positivity rates in the South Korea and Germany were 1% and 1.8%, respectively, during the first wave. The results indicate that the agreement between model predictions and observed data increases as the positive rate decreases. A good agreement between the model predictions and data for weeks ahead for a pandemic region requires an excellent testing capability and a positivity rate under 1% [10].

The results show that both the total number of infections over the course of the pandemic and the daily new infections during the course are sensitive to people’s social behavior as shown in Fig. 3. These numbers increase dramatically when social distancing is relaxed, and the peak time in daily infections is delayed. Reducing social distancing not only increases the length of time until life returns to “normal’’ but also places more lives at risk. This result differs from other studies, which capture the delaying effect of social distancing on the peak number of infections but not the effects on the total and daily number of infections [18, 19]. Stochastic phenomena will emerge due to changes of human behavior. Model predictions are significantly improved by using time dependent social distancing [11]. A model with a stochastic social distancing will be presented in a future paper.

**Fig.3.**
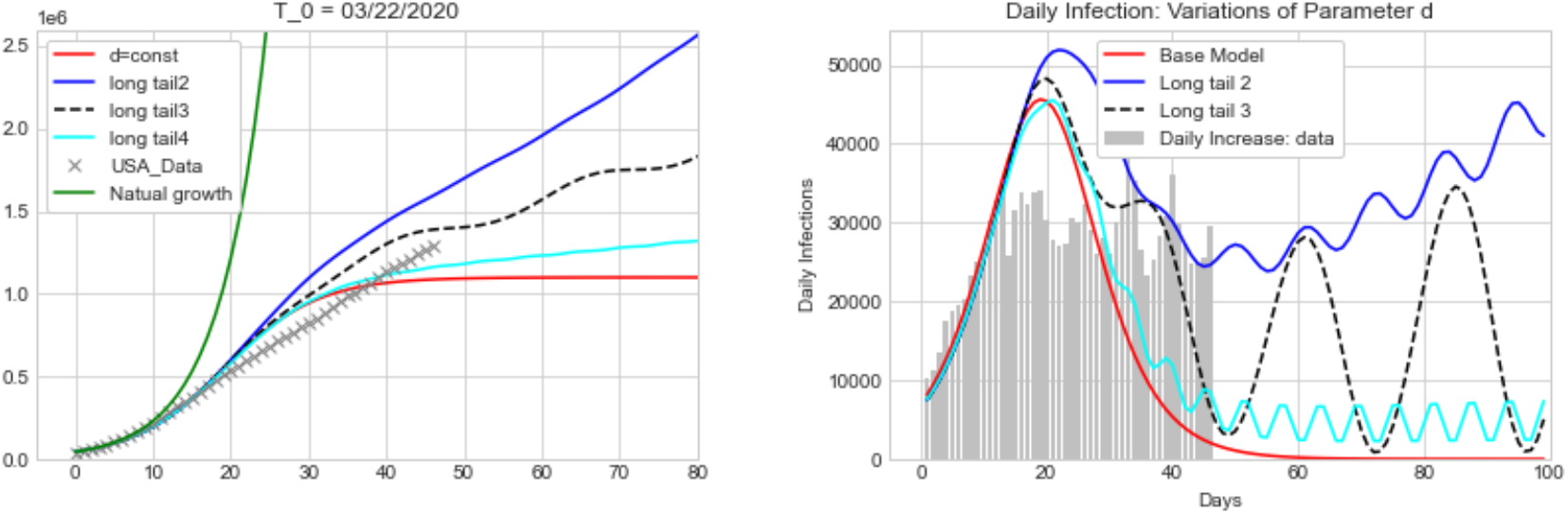
Total number of infections (left) and number of daily new cases (right) changing over time under four social-distancing scenarios. The red line represents the spread trajectory under constant social distancing at the level when restrictions were put in place. The blue line corresponds to social distancing decreasing at a relaxation rate b=1.35 % per day and relaxation period τ=11 days. The black dashed line corresponds to b=0.9% and τ=24 days. The cyan line corresponds to b=0.35% and τ=7 days. All cases assume a community spread rate γ=0.17.

## Conclusion

The newly developed logistic pandemic model has correctly captured the two dynamic phases of the spread of COVID-19, i.e., a natural exponential growth phase that occurs in the absence of intervention and a regulated growth phase that reflects enforcement of social distancing. The two-parameter model is capable to make early predictions about the spreading trajectory in communities of any size including the total infections, the date of peak daily infections and the date of infections reaching a plateau if the testing capability is sufficient. The model has been applied to a number of pandemic centers (during the first wave). The model predictions are in good agreement with the data. The model can place an upper limit for the total number of infections and daily new infections in a pandemic region for weeks into the future, providing the vital information and lead time needed to prepare for and mitigate the pandemic and future waves.

The study shows that the community transmission rate is not uniform across the United States, it depends on the geography and demographics of the infection regions. The uncertainty of the model predictions depends on the quality of data and testing capacity of the pandemic region. The study indicates that most of the US states have been under testing and the data near the peak of daily infections have been significantly saturated by the inability of testing at the time. It is found that effectively controlling and predicting the spread of the COVID-19 pandemic requires a testing at < 1% positivity rate so as to actively tracing and isolating the infections.

The model shows that social distancing has a significant impact in reducing the pandemic spread. Strict social distancing will not only slow the virus spread rate but also significantly reduce the number of total infections, daily infection rates, peak of daily infections, and the duration of the pandemic. Under strict social distancing, the rise and fall of the pandemic are nearly symmetric to the peak time, and the total duration of the pandemic would be around 60 days. If everyone wears a face mask in public, there would not be a need to shut down the economy even if there was a second or third waves of COVID-19. Relaxed social distancing will result in a prolonged pandemic and many more infections and deaths. Recent statistics show that the deaths from flu season in 2020 was significantly less than any other previous years because of the wearing of face masks during the COVID-19 pandemic. This fact supports the evidence of the effectiveness of wearing face masks.

The model exhibits that with the presence of COVID vaccines, the fraction of the population who are immune to the virus (η) increases significantly, so does parameter d_min_. Thus, the pandemic spread will soon be significantly reduced and controlled. Given a percentage of the infected population in a pandemic region, the model could provide a lower limit for the required percentage of vaccinated population in the region so as to control the pandemic. Finally, the vast amount of COVID-19 data collected during the pandemic would provide great opportunities to develop advanced pandemic forecast model and make reliable predictions for weeks ahead using science guided deep learning algorithms, which would have profund applications in the implementation of effective policies for ongoing outbreaks and similar infectious disease pandemic in the future.

## Data Availability

All data are in the public domain as cited in the paper.

## Acknowledgements

The authors are grateful to Dr. P. A. Bradley for useful comments and C. S. Carmer for editing this article. B.C. was supported under the auspices of the U.S. Department of Energy by the Los Alamos National Laboratory under Contract No. 89233218CNA000001.

## Notes

### Competing Interest Statement

The authors have declared no competing interest.

### Author Declarations

Paper has been reviewed by LANL and has a document release number LA-UR-21-22770.

